# Production of Trimeric SARS-CoV-2 Spike Protein by CHO Cells for Serological COVID-19 Testing

**DOI:** 10.1101/2020.08.07.20169441

**Authors:** Yusuf B. Johari, Stephen R.P. Jaffé, Joseph M. Scarrott, Abayomi O. Johnson, Théo Mozzanino, Thilo H. Pohle, Sheetal Maisuria, Amina Bhayat-Cammack, Adam J. Brown, Kang Lan Tee, Philip J. Jackson, Tuck Seng Wong, Mark J. Dickman, Ravishankar Sargur, David C. James

## Abstract

We describe scalable and cost-efficient production of full length, His-tagged SARS-CoV-2 spike glycoprotein trimer by CHO cells that can be used to detect SARS-CoV-2 antibodies in patient sera at high specificity and sensitivity. Transient production of spike in both HEK and CHO cells mediated by PEI was increased significantly (up to 10.9-fold) by a reduction in culture temperature to 32°C to permit extended duration cultures. Based on these data GS-CHO pools stably producing spike trimer under the control of a strong synthetic promoter were cultured in hypothermic conditions with combinations of bioactive small molecules to increase yield of purified spike product 4.9-fold to 53 mg/L. Purification of recombinant spike by Ni-chelate affinity chromatography initially yielded a variety of co-eluting protein impurities identified as host cell derived by mass spectrometry, which were separated from spike trimer using a modified imidazole gradient elution. Purified CHO spike trimer antigen was used in ELISA format to detect IgG antibodies against SARS-CoV-2 in sera from patient cohorts previously tested for viral infection by PCR, including those who had displayed COVID-19 symptoms. The antibody assay, validated to ISO 15189 Medical Laboratories standards, exhibited a specificity of 100% and sensitivity of 92.3%. Our data show that CHO cells are a suitable host for the production of larger quantities of recombinant SARS-CoV-2 trimer which can be used as antigen for mass serological testing.

---

Immune response represents the first line of defense against severe acute respiratory syndrome coronavirus 2 (SARS-CoV-2) infection that has caused the coronavirus disease 2019 (COVID-19) pandemic. The spike glycoprotein that protrudes from the surface of the virus is highly immunogenic with the receptor-binding domain being the target of many neutralizing antibodies (Yuan et al., 2020). Utilizing a stabilized version of the full-length SARS-CoV-2 spike protein, a very robust and accurate serological enzyme-linked immunosorbent assay (ELISA) for antibodies in patient sera has recently been developed (Amanat et al., 2020) and approved for use by the US FDA. However, very low production titers (1-2 mg/L) of the spike trimer were reported using the human embryonic kidney (HEK) Expi293 Expression system (Esposito et al., 2020), therefore effectively limiting its widespread utilization as a preferred antigen in serological assays for COVID-19. The low production titer is not surprising considering that the SARS-CoV-2 spike is a large homotrimer (~600 kDa) with 22 N-linked glycosylation sites per monomer (Watanabe et al., 2020). In this work, we describe the development of spike manufacturing platforms utilizing Chinese hamster ovary (CHO) cells as a preferred production host with established gene amplification methods and improved engineering strategies. Transient expression was initially employed to fast-track the production of CHO spike to perform biophysical analyses and early clinical evaluation of the material, as well as to evaluate product manufacturability and refine process conditions. Even though CHO cells possessed a higher amount of heparan sulfate proteoglycans (HSPGs) compared to HEK cells (Lee et al., 2016) we developed an effective affinity purification procedure, and further validated the CHO-derived spike serological assay for COVID-19 antibody.

We have previously shown that for difficult-to-express (DTE) proteins, transient production processes need to be tailored to negate the protein-specific negative effects of recombinant gene overexpression in host cells (e.g., unfolded protein response (UPR) induction, limited cell-specific productivity (qP); Johari et al., 2015). Using the plasmid construct from the Krammer Laboratory (Amanat et al., 2020), HEK Expi293F and CHO-S cells were transiently transfected with the CAG-driven expression plasmid using PEI at an optimal gene dosage for spike production in both cases (data not shown). Further, we utilized a mild hypothermic condition, an effective process engineering intervention for production of DTE proteins (e.g., Estes et al., 2015; Johari et al., 2015) and to extend culture longevity. As shown in Figure 1A, the qP of HEK cells increased 2.4-fold from 0.20 pg/cell/day to 0.48 pg/cell/day when the culture temperature was lowered from 37°C to 32°C. Additionally, the prolonged batch culture duration at 32°C (Figure 1B) enabled a 4.1-fold increase in titer, yielding 10.2 mg/L of purified spike. Greater enhancement was observed with CHO cells where mild hypothermia resulted in an 8.5-fold higher qP than that at 37°C, and a further increase in titer (10.9-fold, 5.4 mg/L) was obtained via increased cell accumulation (Figure 1A, B). We anticipate that improved CHO systems (e.g., ExpiCHO-S cell line and ExpiCHO medium) in short-term, intensified high-density cell culture maintained at reduced temperature paired with co-expression of genetic effectors and addition of chemical chaperones (Cartwright et al., 2020; Johari et al., 2015; Schmitt et al., 2020) would significantly increase spike transient production in CHO cells. The CHO-derived spike exhibited a monomeric molecular mass of ~190 kDa by SDS-PAGE (Figure 1C) and a trimeric mass of 619 kDa was measured using analytical size exclusion chromatography (Supplementary Figure S1). The material was further validated using peptide mapping in conjunction with mass spectrometry analysis (Supplementary Figure S2, Supplementary Table S1). Critically, the preliminary COVID-19 antibody serological test demonstrated its suitability for the ELISA (data not shown) thus permitting development of CHO stable production platform.

**Figure 1.**
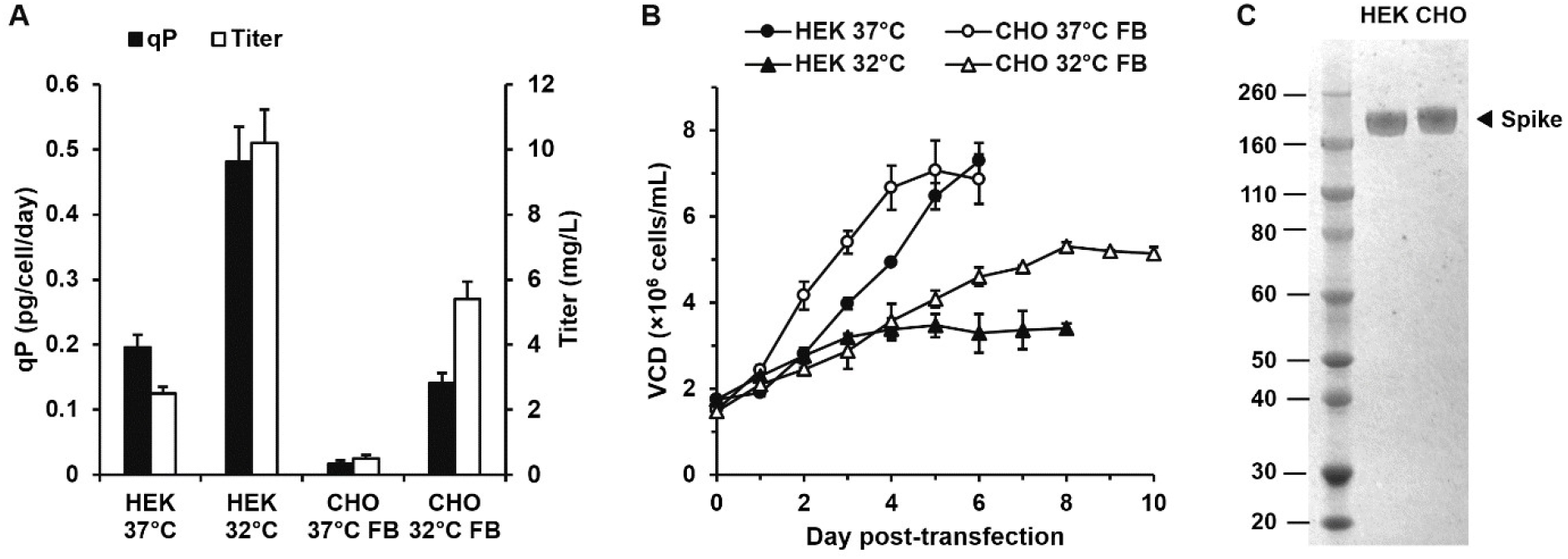
Transient production of recombinant spike in HEK and CHO cells. HEK Expi293F cells and CHO-S cells were transfected with plasmids encoding spike gene using PEI under optimized conditions and cultured at 37°C or 32°C. CHO cultures were fed every 2 days with 5% v/v EfficientFeed B. **(A)** Recombinant qP and titer. **(B)** Viable cell density post-transfection (cell viability >70%). **(C)** Coomassie-stained reducing SDS-PAGE gel of purified HEK and CHO cell-derived spike (~190 kDa).

For DTE proteins, very low yielding transient expression systems can be an early indication of reduced stable production (Mason et al., 2012), where particular engineering strategies may be required to obtain stable cells with desirable production characteristics. To generate CHO cells stably expressing recombinant spike trimer, we tested two in-house CHO synthetic promoters, namely 40RPU (~90% CMV activity) and 100RPU (~220% CMV activity) promoters (see Brown et al., 2017; Johari et al., 2019). Although the use of extremely strong promoters may be counterintuitive for DTE proteins, we reasoned that only those transfectants harboring a sub-UPR threshold productivity, and thus capable of proliferation would survive. Thus, if cell proliferation attenuation and apoptosis occur as ER functional capacity is exceeded, this is a condition that would directly deselect poorly performing stable transfectants. The promoters and spike gene were inserted into a vector construct encoding glutamine synthetase (driven by an SV40 promoter) and the electroporated CHO-S host cells were subjected to a single round of selection at 25 or 50 μM methionine sulfoximine (MSX), using suspension culture. After 19 days, recovered CHO cell populations were screened for the ability to produce spike in 3-day batch culture (Figure 3A). These data showed that transfectant pools derived from genetic constructs harboring the strong 100RPU promoter expressed recombinant spike whereas those using the weaker 40RPU promoter did not. More stringent selection conditions (50 μM MSX) yielded transfectant pools exhibiting lower productivity. Together, these data imply that higher glutamine production may protect cells constitutively expressing DTE spike trimer (e.g. via glutathione production to alleviate oxidative stress). Accordingly, CHO cell pools harboring the 100RPU promoter under 25 μM MSX were taken forward for the manufacturing process.

In order to rapidly produce recombinant spike, stable transfectant pools (rather than clonally derived populations) were employed. Based on CHO transient process data (Figure 1), we tested the hypothesis that an optimal 10-day fed-batch stable production process could be executed at 32°C and further enhanced by chemical chaperone additives (e.g., Johari et al., 2015). We compared this strategy to an alternative approach utilizing reduced culture temperature implemented at maximal cell density (biphasic), as well as constant 37°C as control. These data are shown in Figure 2B and C, whilst the screening data for 9 small molecule chemical additives is shown in Supplementary Figure S3. Compared to the 37°C control culture, hypothermia resulted in a clear (33%) initial reduction in cell-specific proliferation rate over the first 5 days of culture (Figure 2B). However, the qP of the latter was 3.4-fold higher over control and addition of valproic acid (VPA) at Day 6 further enhanced qP 5.1-fold, yielding 51 mg/L of spike after purification by immobilized metal affinity chromatography (IMAC; Figure 2C). Similar enhancement was observed with betaine although there was no synergistic effect when the two molecules were utilized together. Reduction in culture temperature after 3 days culture improved the integral of viable cell density (IVCD) 1.4-fold and when combined with VPA addition at Day 4, 53 mg/L of spike was attained after IMAC purification. These data demonstrate that the optimal process engineering intervention for recombinant spike production identified for rapid transient gene expression was translatable to the stable production process. Based on our data, we speculate that very high stable expression of DTE recombinant spike is inherently not compatible with high cell growth, potentially via induction of an unfolded protein response (UPR). Therefore, low-level, sub-UPR threshold expression is required to permit adequate cell growth. In the transfectant pool, this may be achieved effectively via promoter-mediated transcriptional instability/repression mechanisms such as heterochromatin formation deriving from methylation of CpG islands (Kim et al., 2011). Thus we observe significant de-repression of spike production using HDAC inhibitors in the stable production context. This effectively creates, indirectly, an inducible expression system and suggests that application of mammalian inducible expression technology (e.g. cumate; Poulain et al., 2017) to switch on spike production using an intensified biphasic culture system) would be particularly useful to maximize stable production. We speculate that our use of a lower strength synthetic promoter (40RPU) was therefore theoretically justified, but that clonal derivatives exhibiting significant transcriptional repression of spike production could have outgrown others, rendering spike production undetectable. Whilst repression occurred with clones harboring the high strength promoter (100RPU) also, it’s intrinsically higher transcriptional activity (balanced with an optimal selection stringency) resulted in detectable spike production. Moreover, the synthetic promoters were designed de novo to minimize CpG content (Brown et al. 2017). Of course we also expect that co-expression of product-specific combinations of ER chaperones/UPR regulators may also be a useful strategy to minimise the impact of spike expression on the cell, e.g. by effectively raising the cellular threshold for UPR induction (Johari et al. 2015; Brown et al. 2019; Cartwright et al. 2020).

**Figure 2.**
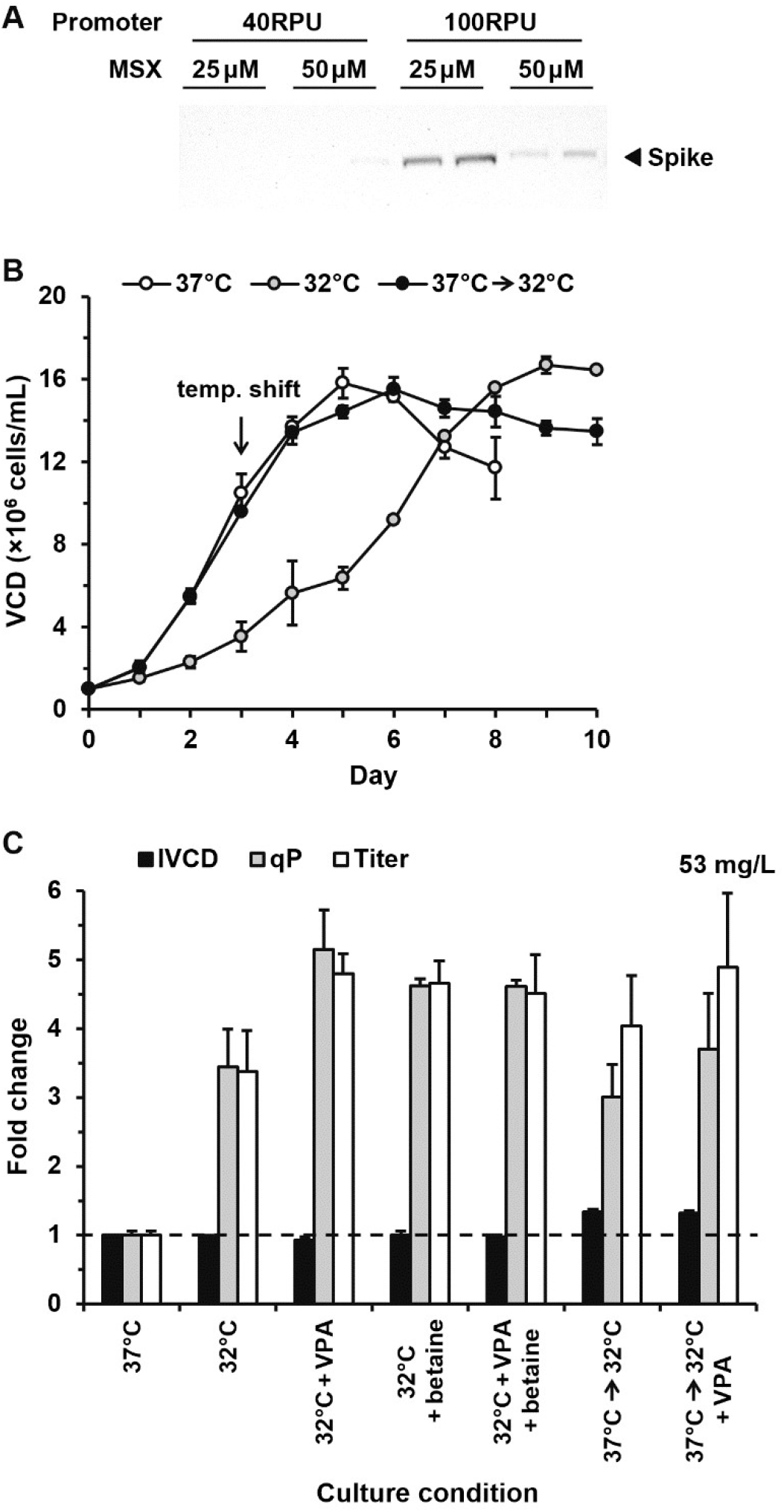
Development of a stable production platform for SARS-CoV-2 spike in CHO cells. **(A)** Generation and analysis of CHO stable transfectant pools expressing recombinant spike under the control of synthetic promoters. CHO-S cells were electroporated in duplicate with plasmids containing a GS gene driven by an SV40 promoter and a spike gene driven by either a 40RPU or 100RPU synthetic promoter, followed by selection in glutamine-free media containing 25 μM or 50 μM MSX under suspension condition. Recovered cell pools were assessed for their ability to express spike in a 3-days batch culture by Western blot. Figure shown is a representative Western blot of two technical replicates. **(B)** Cells from the best performing pools in **A** were inoculated and cultured at 37°C, 32°C, or 37°C with a shift to 32°C at Day 3. Cultures were fed every 2 days with 5% v/v EfficientFeed B. **(C)** Comparison of the fed-batch culture production performance without or with a chemical addition (chemical screening data is shown in Supplementary Figure S2). 1 mM VPA and/or 12.5 mM betaine were added at Day 4 for the biphasic cultures or at Day 6 for the 32°C cultures. Data are normalized with respect to culture at 37°C without any chemical addition. Data shown are the mean value ± standard deviation of two independently generated stable pools each performed in duplicate.

To purify spike protein from culture supernatant, IMAC was initially performed using a step-elution of 250 mM imidazole according to Stadlbauer et al. (2020). Figure 3A shows SDS-PAGE of eluted proteins, and reveals the presence of protein impurities not derived from recombinant spike (Figure 3B), which were identified using tandem mass spectrometry as CHO host cell derived proteins (HCPs; see Supplementary Table S1). Whilst all of the identified extracellular HCPs have previously been shown to be present in CHO cell culture supernatant (Park et al., 2017), HSPG has been reported to occur in CHO cells at relatively higher level than HEK cells (Lee et al., 2016) and shown to interact with spike protein via heparin binding (Mycroft-West et al., 2020). Notably, no ACE2 was identified as a co-eluted protein impurity even though trimeric spike protein tightly binds to this cell surface protein (equilibrium constant, *K*_D_, of 14.7 nM) for cellular entry of the SARS-CoV-2 virus (Lan et al., 2020). The ACE2 receptor has been identified as being highly abundant in human kidney cells and above average in human (Although not necessarily Chinese hamster) ovary cells (Hikmet et al., 2020), albeit in primary tissue instead of derived cell lines. In order to increase recombinant spike purity, a revised gradient elution profile up to 250 mM imidazole was implemented. As shown in Figure 3C, HCPs were eluted at a lower imidazole concentration than recombinant spike, permitting recovery of high purity (>95%) product for use in serological assays (Figure 3D).

**Figure 3.**
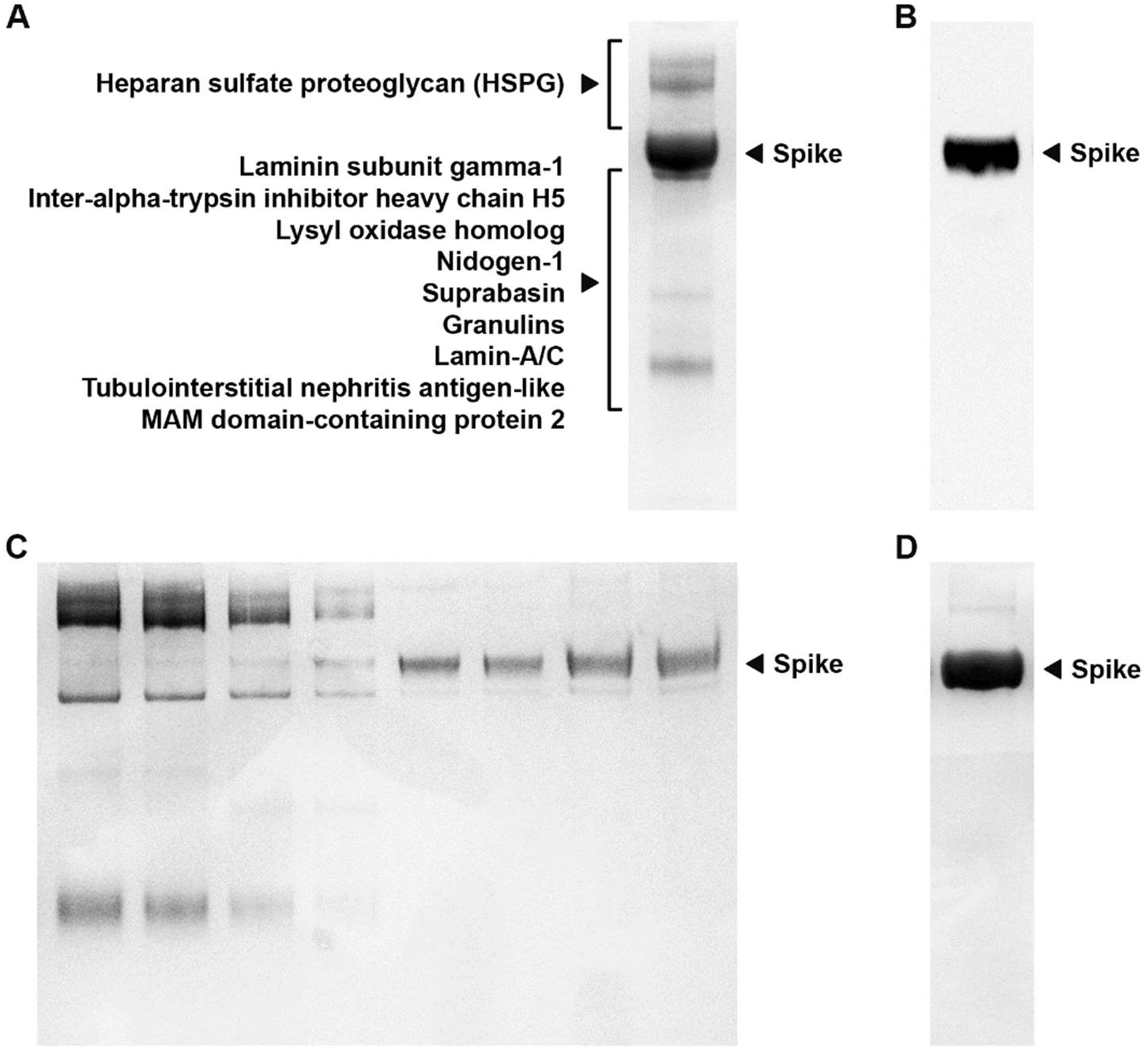
Optimization of affinity chromatography purification strategies for spike protein using HisTrap columns. **(A)** Coomassie stained gel of the initial purification strategy of spike utilizing the method from Stadlbauer et al. (2020) with associated impurities identified using tandem mass spectrometry. **(B)** Assessment of spike sample shown in A by Western Blot. **(C)** Gradient elution of spike protein starting from 10 mM imidazole up to a final concentration of 250 mM imidazole. **(D)** Purified spike from optimized step elution affinity chromatography.

COVID-19 antibody tests would help reveal the true scale of the pandemic in a population and the persistence of immunity, whether vaccines (many of which are based on the production of neutralizing antibodies against spike protein) designed to protect from infection are effective, as well as identify highly reactive human donors for convalescent plasma therapy. The CHO-spike anti-SARS-CoV-2 ELISA was developed based on the Krammer Laboratory’s assay, and validated to ISO 15189 Medical Laboratories standards. Initially, we tested a panel of 234 negative samples taken pre-COVID-19 outbreak (June–August 2019) and 26 positive samples taken during the COVID-19 outbreak (≥15 days post-positive PCR test). ELISAs were performed by 1/20 dilution of the individual serum samples and the cut-off index of 1.4 was determined using the cut-off OD value (ROC curve with 100% specificity) and the negative control. In this particular evaluation, the assay had an overall specificity of 100% and sensitivity of 92.3% as illustrated in Figure 4A. To establish the reproducibility of the ELISA, a positive sample was tested on 5 separate assays over 2 days at 3 different dilutions to determine the inter-assay variations. The data (Figure 4B) shows that the assay performed within the standard range for precision with inter-assay %CV of ≤5%. To be able to interpret serosurveys correctly, the ELISA was evaluated for potential cross-reactivity from individuals with other medical conditions where zero positives were observed in all cases (Supplementary Table S2).

**Figure 4.**
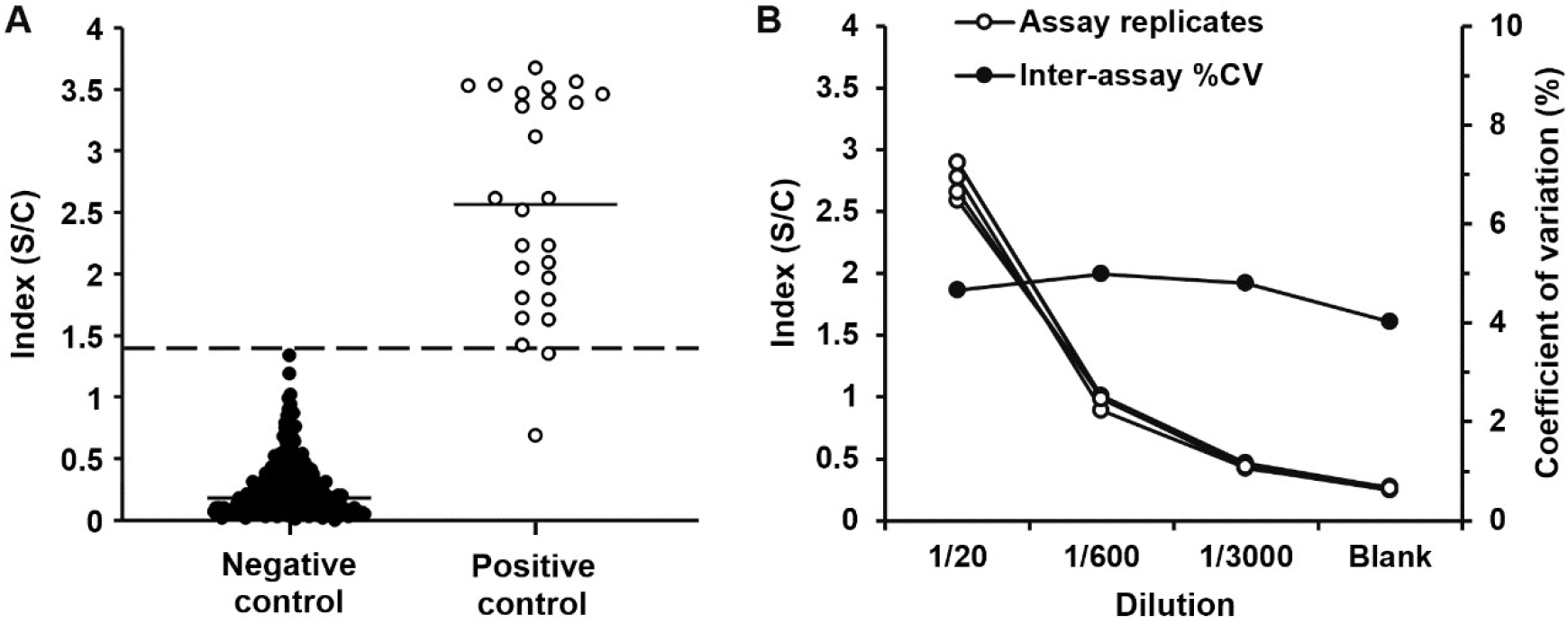
Evaluation of CHO-spike anti-SARS-CoV-2 ELISA. **(A)** 234 negative serum samples (taken pre-COVID-19 outbreak) and 26 positive serum samples (taken ≥15 days post-positive PCR test) were used to evaluate the assay performance, yielding an overall sensitivity of 92.3% anti-SARS-CoV-2 antibodies. **(B)** To determine the assay precision, 1 serum sample was assayed in triplicates at 5 separate times over 2 days (*n* = 15).

Overall, our work serves as an exemplar for a development process of characteristically difficult-to-express spike manufacturing platform utilizing CHO cells. This itself is a significant and useful finding, as many DTE recombinant proteins cannot be produced using this industry standard production vehicle — e.g., a recent study reported that for over 2,200 human genes encoding secreted proteins expressed in CHO cells, almost 50% did not yield target protein (Uhlen et al., 2018). On the other hand, the low spike production in HEK cells was highly dependent on the very expensive Expi293 medium (we note that spike production using FreeStyle 293 medium resulted in an even lower titer (<40%); data not shown). With one mg of spike providing serological assays for approximately 3,500 patient samples, the rapid, scalable transient platform was adequate for local population antibody tests and research studies. To enable large, constant clinical supply of spike, we showed that it was possible to generate CHO stable transfectants expressing the very complex glycoprotein, whilst high titers could be achieved via a combination of process engineering approaches designed for both high qP and cell biomass accumulation. The refinement of the IMAC affinity purification process permitted greatly enhanced purity of the CHO spike product following an extended 10-day culture, ensuring suitability for use in serological immunity testing. The assay has been implemented at local hospitals with ~7,200 staff tested (as of 31 July 2020) which resulted in ~16% positive COVID-19 antibody detection, thus supporting the global effort to limit and mitigate the impact of SARS-CoV-2. Furthermore, it is highly likely that the cell and process engineering interventions designed for SARS-CoV-2 spike production is generically applicable to spike from different coronavirus strains.

## MATERIALS AND METHODS

### Cell Cultures and Chemical Chaperones

Expi293F cells were cultured in Expi293 Expression medium (ThermoFisher) in Erlenmeyer flasks maintained at 37°C, 125 rpm under 8% CO_2_, 85% humidity. CHO-S clonal isolate cells (C1-80; Fernandez-Martell et al., 2018) were cultured in CD CHO medium (ThermoFisher) supplemented with 8 mM L-glutamine maintained at 37°C, 140 rpm under 5% CO_2_, 85% humidity. Cells were seeded at 2×10^5^ viable cells/mL and were sub-cultured every 3–4 days. Cell viability and VCD were measured using the Vi-CELL XR (Beckman Coulter). VPA, NaBu, DMSO, glycerol, betaine, TMAO, proline and FAC were obtained from Sigma while TUDCA was obtained from Merck.

### Transient Production in HEK and CHO Cells

pCAGGS-spike was provided by the Krammer Laboratory (Icahn School of Medicine at Mount Sinai), amplified, and purified using QIAGEN Plasmid *Plus* kit (Qiagen). For Expi293F transfection, cells were grown to 1.75×10^6^ cells/mL, centrifuged and resuspended at a density of 3.5×10^6^ cells/mL, followed by sequential addition of 0.85 μg of DNA and 2.55 μl of PEI MAX (each pre-diluted in 10 μL of 150 mM NaCl) per million cells. At 24 h post-transfection, the cells were diluted 2× by adding fresh medium, and where applicable culture was shifted to 32°C. For CHO transfection, cells were seeded one day before transfection and grown to 1.5×10^6^ cells/mL. For every 1.5×10^6^ cells, 1.3 μg of DNA and 4.55 μL of PEI MAX (each pre-diluted in 15 μL of 150 mM NaCl) were combined and incubated at RT for 2 min before being added into culture. Where applicable, culture was shifted to 32°C at 4 h post-transfection. For fed-batch production, 5% v/v CHO CD EfficientFeed B (ThermoFisher) was added at Days 2, 4, 6 and 8.

### Generation of Stable CHO Pools and Fed-batch Production

A stable vector containing an SV40 promoter-driven GS gene was provided by AstraZeneca, UK. The spike gene was cloned by PCR, inserted into the vector downstream of 40RPU or 100RPU synthetic promoter (Brown et al., 2017) and the plasmid constructs were confirmed by DNA sequencing. 10×10^6^ cells per cuvette were electroporated with 7 μg linearized DNA using Cell Line Nucleofector Kit V system (Lonza) and transferred to a TubeSpin containing 10 mL glutamine-free culture medium with the addition of 25 or 50 μM MSX after 48 h. The cells were left to recover under suspension conditions and recovered pools were cryopreserved when the cell viability reached >90%. For fed-batch production, 5% v/v CHO CD EfficientFeed B was added at Days 2, 4, 6 and 8.

### Western Blotting

Proteins in culture supernatant were precipitated by TCA/DOC, resuspended in LDS loading buffer with BME and heated to 70°C. SDS-PAGE was performed using 4–12% NuPAGE Bistris gels and resolved proteins were transferred to nitrocellulose membranes by iBlot system (ThermoFisher). Membranes were blocked in 5% milk/TBS-T before being incubated with HRP-conjugated anti-HisTag antibody (Bio-Rad) and visualized by enhanced chemiluminescence (ECL; ThermoFisher).

### Recombinant Protein Purification and Quantification

Spike protein was harvested by centrifugation at 3,000×g for 20 min at 4°C and supernatant was filtered through a 0.22 μm filter. Protein was purified using the ÄKTA Pure system (Cytiva) and a 5-mL HisTrap HP column (Cytiva). Eluted protein fractions were pooled and buffer exchanged into storage buffer (20 mM Tris, 200 mM NaCl, 10% v/v glycerol, pH 8.0) using a PD-10 desalting column (Cytiva). Protein was quantified using the Pierce Coomassie Plus (Bradford) Assay kit (ThermoFisher) and analyzed by reducing SDS-PAGE. A complementary quantification of spike in culture supernatant was performed using CR3022 antibody ELISA (Supplementary Figure S4).

### Protein Identification by Mass Spectrometry

All materials were supplied by ThermoFisher unless otherwise stated. Briefly, protein samples in 50 mM ammonium bicarbonate (ABC), 5 mM tris(2-carboxyethyl)phosphine-HCl were reduced by incubation at 37°C for 30 min. S-alkylation was performed by the addition of 1 μL 100 mM methyl methanethiosulfonate in isopropanol. For proteolytic digestion, 1.5 μL 0.2% ProteaseMax surfactant in 50 mM ABC and 2 μL 0.2g/L trypsin/endoproteinase Lys-C mixture (Promega) were added followed by incubation at 37°C for 16 h. Proteolysis was stopped and the surfactant hydrolyzed by the addition of 0.5% trifluoroacetic acid (TFA). The samples were desalted using HyperSep Hypercarb solid phase extraction tips and dried by vacuum centrifugation. For RPLC-MS, samples in 0.5% TFA, 3% acetonitrile (ACN) were injected. Peptides were separated using an RSLCnano system with a PepSwift PS-DVB monolithic column using a gradient from 97% solvent A (0.1% formic acid) to 35% solvent B (0.1% formic acid, 80% ACN). Mass spectra were acquired on a Q Exactive HF quadrupole-Orbitrap instrument, with automated data dependent switching between full-MS and tandem MS/MS scans. Proteins were identified by converting the MS data into Mascot Generic Format (MGF) files and analyzed against human and Chinese hamster reference proteome databases with the spike glycoprotein construct sequence inserted (www.uniprot.org) using Mascot Daemon v.2.5.1 with Mascot server v.2.5 (Matrix Science). Search parameters and protein identifications are detailed in Supplementary Table S1.

### Spike ELISA

The ELISA protocol was adapted from Stadlbauer et al. (2020) using spike protein with >95% purity. Microtiter plates (96-well) were coated overnight with 50 μL of spike per well (2 μg/mL in PBS pH 7.4) at 4°C. The coating solution was removed and 300 μL of blocking solution (3% non-fat milk in 0.1% PBS-T) was added for 1 h and washed 3 times (with 0.1% PBS-T). Samples were added at 1/20 dilution and incubated for 2 h at RT. Plate was washed 3 times and 100 μL of anti-human IgG conjugate was added to the wells and incubated for 1 h at RT. Plate was washed 3 times and 100 μL of substrate was added and incubated in the dark for 45 minutes. The reaction was stopped by the addition of 50 of μl 3 M HCl and the plate was read at 490 nm using the Agility (Dynex Technologies) ELISA system. The index value was calculated as follows:

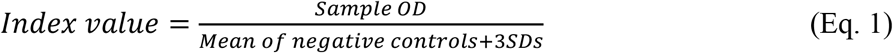

The cut-off value was calculated with 100% specificity using ROC curves of calculated index values.

## Data Availability

All data discussed in the text is submitted as supplementary information.

## ACKNOWLEDGEMENTS

This work was supported by Sheffield Teaching Hospitals NHS Foundation Trust, UK. The authors are grateful to Prof. Florian Krammer (Icahn School of Medicine at Mount Sinai, New York) for providing the spike plasmid via Dr. Thushan de Silva (University of Sheffield), Martin Nicklin (University of Sheffield) for providing the Expi293F cells, Molly Smith (University of Sheffield) for a preliminary test of HEK transfection procedures, Prof. William Egner (Sheffield Teaching Hospitals) for support and help with ELISA development work and validation. MJD acknowledges support from the Biotechnology and Biological Sciences Research Council UK (BBSRC) (BB/M012166/1).

